# Malaria parasite prevalence in asymptomatic migrants residing in Sweden

**DOI:** 10.1101/2022.09.09.22279774

**Authors:** Andreas Wångdahl, Rebecca Tafesse Bogale, Isabelle Eliasson, Ioanna Broumou, Fariba Faroogh, Filip Lind, Ganna Vashchuk, Adina Hildell, Suzanne Franson, Emil Hallberg, Isabelle Grip, Irene Nordling, Angelica Gervin, Shelan Kaitoly, Berhane Tekleab, Katja Wyss, Ana Requena Mendez, Olof Hertting, Anna Färnert

## Abstract

**Background:** Asymptomatic infections with malaria parasites are common in populations in endemic areas. These infections may persist in migrants after arrival in a non-endemic area. Screening to find and clear these infections is generally not implemented in non-endemic countries, despite a potential negative health impact. We performed a study to evaluate the *Plasmodium* parasite prevalence in migrants living in Sweden.

**Methods:** Adults and children born in Sub-Saharan Africa (SSA) were invited in the study between April 2019 and June 2022 at 10 different sites. Rapid diagnostic tests (RDT) and real-time PCR were used to detect malaria parasites. Prevalence and test sensitivity were calculated with 95% confidence intervals (CI). Univariate and multivariable logistic regression were used to evaluate associations with PCR positivity.

**Findings:** In total, 789 individuals were screened for *Plasmodium spp*., of which 71 (9·0%) were detected by PCR and 18 (2·3%) were also RDT positive. A high prevalence was detected in migrants with Uganda as the country of last residence, 53/187 (28·3%), particularly in children, 29/81 (35·8%). Among the PCR positive, 47/71 (66·2%) belonged to families with at least one other member testing positive (OR 43·4 (95% CI 19·0-98·9), and the time lived in Sweden ranged between 6-386 days.

**Interpretation:** A high malaria parasite prevalence was found in migrants from SSA, particularly in children. Awareness of asymptomatic malaria infection is needed and screening for malaria in migrants arriving from high endemic countries should be considered.

**Funding:** Swedish Research Council, Stockholm County Council and Centre for Clinical Research, Västmanland, Sweden.

**Research in context:** *Evidence before this study:* Asymptomatic malaria infections are common in endemic areas, and migrants may still be infected when arriving in a non-endemic country. Previous studies have shown a parasite prevalence between 3-31.3% in migrants arriving in non-endemic countries, largely depending on patient origins and the diagnostic method used. No European country recommends screening for malaria, whereas in Australia screening of high risk groups is recommended, and in the US, presumptive antimalarial treatment is given. Robust data are few to establish which migrants are at highest risk of infection and who should be screened. In addition, the duration of asymptomatic plasmodium infections has not been assessed thoroughly, and available data are based on case-series which may represent extreme examples of long duration.

*Added value of this study:* This study is one of the largest cross-sectional studies that evaluate the prevalence of malaria in migrants living in a non-endemic country. The study was primarily conducted in a primary health care level, resembling a potential way to screen for malaria and to avoid the overestimation of the prevalence which is usually perceived in hospital-based studies. Apart from describing a parasite prevalence of 9% in asymptomatic migrants, using real-time PCR, we also identified country of last residence as one of the key indicators to identify the risk of carrying malaria parasites, with a parasite prevalence of 53/187 (28.3%) in individuals where Uganda was the country of last residence. Other risk factors for PCR-positivity were evaluated and children were at particular risk of PCR positivity clustering of asymptomatic malaria infections in families are described. Duration of infection could also be estimated due to the lack of re-exposure for malaria in Sweden.

*Implications of all the available evidence:* The results presented in this study summarise the best available epidemiological information for the prevalence of malaria according to PCR and RDT techniques among a large sample of migrants living in a non-endemic setting and can be used to inform screening decisions, suggesting that screening for malaria is motivated in migrants from Sub-Saharan Africa after arrival in a non-endemic country.

## Background

Malaria is a major threat to public health globally, especially in Sub-Saharan Africa.^1^ In populations living in endemic areas, acquired immunity gradually evolves over the course of repeated infections. The immunity may eventually control the infection resulting in asymptomatic malaria infections, in some areas affecting more than 50% of the population and is recognised as a reservoir for transmission.^2^

Migrant populations in Sub-Saharan Africa (SSA) countries have been reported to be at particular risk of malaria,^3^ and may have persistent infections after arrival in non-endemic countries.^4-7^ Previous studies on migrants arriving in non-endemic countries have shown a malaria parasite prevalence between 3% in all-origin migrants in Canada,^5^ to 31·8% in migrants from Africa in an Italian refugee camp,^4^ illustrating that the prevalence may vary between receiver countries, due to differences in the origin of migrant populations, and over time.^4,5,7,8^ Migrant groups at particular risk for malaria infection remain to be identified. In addition, the duration of parasite carriage has not been fully elucidated and present data relies on older data and case series.^9^

Apparently asymptomatic malaria infections may have negative consequences, and are well recognised to cause anaemia, increased risk of serious adverse events during pregnancy, such as maternal and infant mortality, spontaneous abortion, and low birthweight,^10^ hyperreactive malarial splenomegaly,^11^ and have been associated with cognitive dysfunction, bacterial infections and all-cause mortality.^12^

In countries with high and moderate transmission, the World Health Organization (WHO) recommends intermittent preventive treatment of malaria during pregnancy (IPTp) and for infants (IPTi).^13^ However, migrant populations are unlikely to be reached by these measures after arriving in non-endemic countries, despite the risk of continuous parasitemia. In Sweden, quota refugees and asylum seekers are offered a free health assessment after arrival. Testing for malaria is evident in febrile patients from endemic countries, but malaria is not part of the routine health screening for migrants in Sweden. Malaria is not mentioned in the European Centre for Disease Prevention and Control (ECDC) guidance on infectious diseases screening of newly arrived migrants,^14^ while the Australasian Society for Infectious Diseases recommend screening for malaria in migrants from endemic areas,^15^ and in the US, the Centers for Disease Control and Prevention (CDC) recommends presumptive treatment (i.e., without testing) with artemether/lumefantrine for migrants arriving from Sub-Saharan Africa.^16^

Estimating the burden of malaria in newly arrived migrants and in immigrants with longer residence in non-endemic countries could inform strategies for improved migrant health and increase our knowledge about duration of asymptomatic infections in the absence of re-exposure. The study aimed to assess malaria parasite prevalence in migrants from SSA with different duration of residence in Sweden.

## Materials and methods

### Study design and study population

This cross-sectional study was carried out between April 2019 and June 2022. Individuals born in Sub-Saharan Africa were invited to participate, irrespective of length of residence in Sweden. All participants, or their legal guardian, were informed about the study and provided written informed consent with translator assistance. Participants were recruited in six different ways. First, participation in the study was offered adults and children attending a Migrant Health Clinic (Primary Health Clinics in Rissne, Fittja and Skärholmen) and Västerås. Second, the Antenatal care facility at Rissne included pregnant women from SSA. Third, patients of SSA origin attending the Infectious Diseases or Paediatric out-patient clinic at Karolinska University Hospital in Stockholm were invited when at a regular visit for other causes, e.g. latent tuberculosis treatment. Fourth, migrants with origin in Uganda and Democratic Republic of Congo (DRC) living in Stockholm with arrival between 1 January 2015 and 1 October 2022, were contacted with an invitation letter, based on an interim analysis in May 2020 showing PCR positivity in this group. The addresses were acquired from the Swedish Migration Agency. Fifth, community advertisement with posters and presentations at an arranged event and in language classes were performed in 2022. Lastly, individuals with SSA origin related to patients with confirmed malaria, either in the study or as clinical cases at Karolinska University Hospital were invited to participate.

Participants with positive rapid diagnostic test for malaria (RDT) or polymerase chain reaction (PCR) were referred to the Karolinska University Hospital for clinical evaluation and antimalarial treatment.

### Data collection and sampling

A questionnaire was filled by a study team member, collecting information on participants’ origin, route of migration, date of arrival in Sweden, co-morbidities and medications, previous antimalarial treatments, and ongoing symptoms. A peripheral blood sample was collected in an ethylenediaminetetraacetic acid (EDTA) tube. Haemoglobin was measured using a point-of-care HemoCue Hb201 (HemoCue, Sweden).

### Detection of malaria parasites

Rapid diagnostic test (RDT) for malaria (antigen detection test CareStart Malaria HRP2/pLDH, Access Bio, Somerset, NJ, USA) was performed on the sampling day. After centrifugation, DNA-extraction was done on the blood cell fraction using QIAmp DNA Blood Mini Kit (Qiagen, Germany) according to the manufacturer’s manual. A multiplex real-time PCR assay detecting the 18S gene of *Plasmodium falciparum, P. vivax, P. ovale* and *P. malariae*, was performed as previously described by Shokoples et al.^17^ All samples were run in plate duplicates.

### Definitions

Anaemia was defined according to the WHO: haemoglobin concentration <110g/L in ages 0-5 years and during pregnancy, <115g/L in ages 5-11 years, <120 g/L in ages 12-14 years and in non-pregnant adult women, and <130 g/L in men aged >15 years. Severe anaemia was defined as haemoglobin <80g/L, or <70g/L in children under 5 and during pregnancy.^18^

### Statistical analyses

Numerical variables were analysed by Mann-Whitney’s U-test, whereas categorical variables were compared using Pearson’s chi-squared test or Fisher test when appropriate. Prevalence and test sensitivity were calculated with 95% confidence intervals (CI). Univariate and multivariable logistic regression, applying cluster robust standard errors, was used to determine risk factors contributing to PCR positivity, expressed as odds ratio (OR) and adjusted OR (aOR) with 95% CI, adjusting for age group, sex, time in Sweden and previous antimalarial treatment within the past year, as these factors were regarded as clinically relevant. Maximum likelihood ratio test was used to evaluate the model. P-values ≤0.05 were regarded as statistically significant. Analyses were performed in Stata version 14.2 (StataCorp, College Station, TX, USA).

### Ethical considerations

The study was approved by the Swedish Ethical Review Authority, (2019-00430 with amendment 2020-05351). The study was registered at Clinical Trials (NCT05086887). All study procedures agreed with the Helsinki Declaration.

## Results

In total, 789 study participants born in Sub-Saharan Africa were included in the study; 480 participants at the Migrant Health Clinics, 199 from the Infectious Diseases or Paediatric outpatient clinics at Karolinska University Hospital, 54 responders to 277 invitation letters sent, 28 responders to community announcements, 15 relatives to individuals with confirmed malaria infection, and 13 participants from the Antenatal Health Clinic in Rissne. Characteristics of the study population are presented in Table 1.

**Table 1.** Characteristics of the study population.

| n (%) | Site of inclusion |  |  |  |  |  |  |
| --- | --- | --- | --- | --- | --- | --- | --- |
|  | All | Migrant Health Clinics | Antenatal Health Clinic | Infectious Diseases or Paediatric Clinics | Letter invitations to migrants from DRC or Uganda | Community advertisement | Family screening around index cases with confirmed malaria |
| **All** | 789 | 480 (60.8) | 13 (1.6) | 199 (25.2) | 54 (6.8) | 28 (3.6) | 15 (1.9) |
| **Age group** |  |  |  |  |  |  |  |
| 0-5 | 56 (7.1) | 47 (9.8) | - | - | 3 (5.6) | - | 6 (40.0) |
| 6-12 | 95 (12.1) | 81 (16.9) | - | 1 (0.5) | 7 (13.0) | 3 (10.7) | 3 (20.0) |
| 13-18 | 125 (15.8) | 101 (21.0) | - | 10 (5.0) | 11 (20.4) | 2 (7.1) | 1 (6.7) |
| 19-39 | 369 (46.8) | 189 (39.4) | 12 (92.3) | 137 (68.9) | 18 (33.3) | 9 (32.1) | 4 (26.6) |
| 40-59 | 109 (13.8) | 43 (9.0) | 1 (7.7) | 39 (19.6) | 13 (24.1) | 12 (42.9) | 1 (6.7) |
| ≥60 | 27 (3.4) | 13 (2.7) | - | 12 (6.0) | 1 (1.8) | 1 (3.6) | - |
| Missing data | 8 (1.0) | 6 (1.2) | - | - | 1 (1.8) | 1 (3.6) | - |
| **Sex** |  |  |  |  |  |  |  |
| Male | 380 (48.2) | 236 (49.2) | - | 96 (48.2) | 30 (55.6) | 11 (39.3) | 7 (53.3) |
| Female | 409 (51.8) | 244 (50.8) | 13 (100) | 103 (51.8) | 24 (44.4) | 17 (60.7) | 8 (46.7) |
| *Pregnancy (% of women in fertile age)* | 39 (14.8) | 6 (4.4) | 13 (100) | 19 (20.2) | - | - | 1 (33.3) |
| **Country of birth** |  |  |  |  |  |  |  |
| Congo (DRC and RC) | 189 (24.0) | 123 (25.6) | - | 18 (9.1) | 41 (75.9) | 1 (3.6) | 6 (40.0) |
| Eritrea | 156 (19.8) | 90 (18.8) | 6 (46.2) | 52 (26.1) | - | 8 (28.6) | - |
| Sudan and South Sudan | 86 (10.9) | 64 (13.3) | - | 16 (8.0) | - | 6 (21.4) | - |
| Somalia | 97 (12.3) | 28 (7.1) | 1 (7.6) | 44 (22.1) | - | 7 (25.0) | - |
| Uganda | 60 (7.6) | 36 (7.5) | - | 5 (2.5) | 13 (24.1) | - | 6 (40.0) |
| Other<sup>a</sup> | 201 (25.4) | 139 (29.0) | 6 (46.2) | 64 (32.2) | - | 6 (21.4) | 3 (20.0) |
| **Last Sub-Saharan African country of residence** |  |  |  |  |  |  |  |
| Uganda | 187 (23.7) | 129 (26.9) | - | 21 (10.5) | 27 (50.0) | - | 10 (66.7) |
| Ethiopia | 92 (11.7) | 57 (11.9) | 2 (15.4) | 28 (14.1) | 1 (1.9) | 2 (7.1) | - |
| Somalia | 65 (8.2) | 30 (6.3) | - | 32 (16.1) | - | 3 (10.7) | - |
| Sudan and South Sudan | 62 (7.8) | 43 (9.0) | - | 14 (7.0) | - | 4 (14.3) | - |
| Rwanda | 40 (5.1) | 35 (7.3) | - | 1 (0.5) | 3 (5.6) | 1 (3.6) | - |
| Other<sup>b</sup> | 343<br>(43.5)<sup>c</sup> | 186 (38.8) | 11 (84.6) | 103 (51.8) | 23 (42.6) | 18 (64.3) | 5 (33.3) |
| **Time in Sweden** |  |  |  |  |  |  |  |
| **Median, days (range)** | 117 (5-21170) | 56 (5-5353) | 2373 (274-5110) | 730 (9-21170) | 337 (57-4380) | 2931 (13-13608) | 329 (72-425) |
| **Days** |  |  |  |  |  |  |  |
| **0-30** | 120 (15.2) | 112 (23.3) | - | 2 (1.0) | - | 6 (21.4) | - |
| **31-90** | 209 (26.5) | 197 (41.1) | - | 10 (5.0) | 1 (1.9) | - | 1 (6.7) |
| **91-180** | 102 (12.9) | 75 (15.6) | - | 24 (12.1) | - | - | 3 (20.0) |
| **181-365** | 113 (14.3) | 37 (7.7) | 1 (7.7) | 34 (17.1) | 33 (61.1) | - | 8 (53.3) |
| **366-550** | 41 (5.2) | 15 (3.1) | - | 14 (7.1) | 8 (14.8) | 1 (3.6) | 3 (20.0) |
| **551-730** | 14 (1.8) | 1 (0.2) | 1 (7.7) | 12 (6.0) | - | - | - |
| **≥731** | 150 (19.0) | 12 (2.5) | 11 (84.6) | 95 (47.7) | 12 (22.2) | 20 (71.4) | - |
| **Missing data** | 40 (5.1) | 31 (6.5) | - | 8 (4.0) | - | 1 (3.6) | - |
| **Reported ongoing symptoms<sup>c</sup>** | 80 (10.1) | 45 (9.4) | - | 21 (10.6) | 10 (18.5) | 1 (3.6) | 3 (20.0) |
| **Reported fever last month** | 103 (13.3) | 45 (12.7) | - | 47 (35.6) | 3 (9.4) | 4 (14.3) | 4 (26.7) |
| **Anaemia<sup>d</sup>** | 153 (19.4) | 98 (20.4) | 5 (38.5) | 34 (17.1) | 6 (11.1) | 8 (28.6) | 2 (13.3) |
| **Reported previous antimalarial treatment, within 12 months** | 66 (8.4) | 59 (12.3) | - | 5 (2.5) | - | - | 2 (13.3) |

Abbreviations: DRC – Democratic Republic of the Congo. RC – Republic of the Congo. CAR – Central African Republic
<sup>a</sup> Other: Burundi (4), CAR (6), Cameroon (4), Djibouti (2) Eswatini (3), Ethiopia (65), Gambia (8), Ghana (5), Ivory Coast (4), Kenya (9), Liberia (1), Malawi (3), Mali (2), Mozambique (1), Nigeria (10), Rwanda (27), Senegal (3), Sierra Leone (1), Tanzania (16), Zambia (22), Zimbabwe (4), Missing data (1)
<sup>b</sup> Other Sub-Saharan country: Burundi (4), Cameroon (4), CAR (4) Eritrea (27), Eswatini (7), Gambia (6), Ghana (4), Guinea (1), Ivory Coast (2), Kenya (31), Malawi (26), Mali (2), Mauretania (2) Niger (35), Nigeria (11), Senegal (4), Sierra Leone (1), South Africa (4), Tanzania (21), Zambia (40), Missing data (107)
<sup>c</sup> Headache (n=29), stomach-ache (18), cough (8), other (25), confirmed fever ≥38.0°C (3)
<sup>d</sup> Anaemia was defined according to the WHO definition, Haemoglobin <110g/L in ages 0-5 years, <115g/L in ages 5-11 years, <120 g/L in ages 12-14 years, <120 g/L in non-pregnant adult women and <110 g/L in pregnant women, and <130 g/L in men aged >15 years. Severe anaemia was defined as haemoglobin <80g/L, except in children under 5 and during pregnancy when instead <70g/L was the cut-of (25).

### Plasmodium detection by RDT and real-time PCR

PCR for *Plasmodium sp*. was positive in 71/789 (9·0%, 95% CI 7·1-11·2), whereas RDT was positive in 18/789 (2·3%, 95% CI 1·4-3·6). In participants recruited at the Migrant Health Clinic, representing an unselected group of migrants from SSA, PCR positivity for *Plasmodium sp*. was found in 50/480 (10·4%), and RDT was positive in 16/480 (3·3%). The prevalence in migrants included as part of screening of relatives to confirmed malaria cases was 10/15 (66·7%) by PCR and 1/15 (6·7%) by RDT, and among letter invited with origin in DRC or Uganda 10/54 (18·5%) and 1/54 (1·9%), respectively. Only one PCR positive participant was seen among patients attending the out-patient clinics, 1/199 (0.5%), and RDT was negative. This was an adult patient originating from DRC living in Sweden for 386 days when tested.

All RDT positive samples in the study were PCR positive. The overall sensitivity of RDT compared to PCR was 25·4% (95% CI 15·8-37·1) (Supplementary table 1). All four *Plasmodium* species were identified by PCR (Table 2).

**Table 2.** Host factors and *Plasmodium sp*. detected by PCR.

| **n (prevalence, %)** | ***P. falciparum*** | ***P. ovale*** | ***P. malariae*** | **Mixed species<sup>a</sup>** | **All species prevalence, % (95% CI)** |
| --- | --- | --- | --- | --- | --- |
| **All** | 35 (4.4) | 17 (2.2) | 8 (1.0) | 11 (1.4) | 9.0 (7.1-11.2) |
| **Age group** |  |  |  |  |  |
| 0-5 | 5 (8.9) | 8 (14.3) | 1 (1.8) | - | 25.0 (14.4-38.4) |
| 6-12 | 7 (7.4) | 3 (3.2) | 2 (2.1) | 2 (2.1) | 14.7 (8.3-23.5) |
| 13-18 | 6 (4.8) | 1 (0.8) | 1 (0.8) | 3 (2.4) | 8.8 (4.5-15.2) |
| 19-39 | 13 (3.5) | 3 (0.8) | 2 (0.5) | 4 (1.1) | 6.0 (3.8-8.9) |
| 40-59 | 4 (3.7) | 2 (1.8) | 1 (0.9) | 2 (1.8) | 9.3 (4.1-17.5) |
| ≥60 | - | - | 1 (3.7) | - | - |
| Missing data | - | - | - | - |  |
| **Sex** |  |  |  |  |  |
| Male | 20 (5.3) | 7 (1.9) | 1 (0.3) | 4 (1.1) | 8.4 (5.8-11.7) |
| Female | 15 (3.7) | 10 (2.4) | 7 (1.7) | 7 (1.7) | 9.5 (6.9-12.8) |
| *Pregnancy* | 1 (2.6) | 0 | 0 | 0 | 2.6 (0.1-13.5) |
| **Duration of residence in Sweden, days** |  |  |  |  |  |
| Median (range) | 94 (6-386) | 28 (7-371) | 90.5 (23-356) | 134 (27-355) |  |
| <90 | 17 (5.2) | 12 (3.7) | 4 (1.2) | 5 (1.5) | 11.6 (8.3-15.5) |
| ≥90 | 17 (4.1) | 5 (1.2) | 4 (1.0) | 6 (1.4) | 7.6 (5.3-10.6) |
| <365 | 32 (6.0) | 16 (3.0) | 8 (1.5) | 11 (2.0) | 10.8 (8.2-13.9) |
| ≥365 | 3 (1.2) | 1 (0.4) | - | - | 1.7 (0.3-4.9) |
| Missing data | 1 (2.5) | - | - | - |  |
| ***In migrants arrived from Uganda only, days in Sweden*** |  |  |  |  |  |
| <90 | 11 (15.1) | 12 (10.5) | 4 (3.5) | 5 (4.4) | 22.7 (15.5-31.6) |
| ≥90 | 9 (7.9) | 3 (4.1) | 3 (4.1) | 6 (8.2) | 27.6 (17.4-39.6) |
| <365 | 18 (10.8) | 14 (8.4) | 7 (4.2) | 11 (6.6) | 12.5 (9.8-15.5) |
| >365 | 2 (11.1) | 1 (5.0) | - | - | 1.6 (0.06-4.2) |
| **Reported ongoing symptoms<sup>b</sup>** | - | 4 (5.0) | 2 (2.5) | 1 (1.3) | 3.4 (0.1-17.8) |
| **Reported fever, ongoing or in the last month** | 1 (1.0) | 5 (4.9) | 2 (2.0) | 2 (1.9) | 9.7 (4.8-17.1) |
| **Reported previous antimalarial treatment, within 12 months** | 5 (7.6) | 5 (7.6) | 3 (4.6) | - | 19.7 (10.9-31.3) |
| **Anaemia<sup>c</sup>** | 8 (5.2) | 7 (4.6) | 1 (0.7) | 4 (2.6) | 13.1 (8.2-19.5) |
| **Country of last residence** |  |  |  |  |  |
| Uganda | 20 (10.7) | 15 (8.0) | 7 (3.7) | 11 (5.9) | 28.3 (22.0-35.4) |
| Tanzania | 5 (23·8) | 1 (4·8) | - | - | 28·6 (11·3-52·2) |
| Niger | 3 (8·8) |  |  |  | 8·8 |
| Zambia | 2 (5·0) | - | - | - | 5·0 (0·6-16·9) |
| CAR | - | 1 (25·0) | 1 (25·0) | - | 50·0 (0·6-93·2) |
| Congo (DRC and RC) | 2 (13·3) | - | - | - | 13·3 (0·2-40·5) |
| Rwanda | 1 (2·5) | - | - | - | 2·5 (0·06-13·2) |
| Cameron | 1 (25·0) | - | - | - | 25 (0·6-80·1) |
| Malawi | 1 (3·8) | - | - | - | 3·8 (0·1-19·6) |

Abbreviations: PCR – Polymerase Chain Reaction, CI – Confidence interval, *P.* – *Plasmodium*, CAR – Central African Republic, DRC – Democratic Republic of the Congo, RC – Republic of the Congo.
<sup>a</sup> Mixed infection consisted of *P. falciparum* and *P. ovale* (3), *P. falciparum* and *P. malariae* (3), *P. vivax* and *P. malariae* (2), *P. ovale* and *P. malariae* (1), *P. falciparum*, *P. ovale* and *P. malariae* (2), and *P. falciparum*, *P. vivax*, *P. ovale* and *P. malariae* (1)
<sup>b</sup> Reported symptoms consisted of cough (3) or body pain (1) in *P. ovale*, cough (1) or body pain (1) in *P. malariae* and headache (1) in the mixed *Plasmodium* infection.
<sup>c</sup> According to WHO, see table 1 for definition (25).

### Host factors associated with PCR positivity

Among the 71 PCR positive individuals, 53/71 (74·7%) had resided in Uganda before arrival in Sweden. Among the PCR positive migrants arriving from Uganda, 27/53 (50·9%) were born in DRC, and another 12/53 (22·6%) were born in Uganda but had parents originating in DRC. In addition, PCR positivity was also found in individuals arriving from Tanzania 6/21 (28·6%), Niger 3/35 (8·6%), Central African Republic 2/4 (50·0%), DRC 2/15 (13·3%), Zambia 2/30 (6·7%), Cameroon 1/7 (42·9%), Malawi 1/26 (3·8%), and Rwanda 1/38 (2·6%).

Among migrants with Uganda as the country of last residence, the prevalence was 53/187 (28·3%). Living in Uganda before arrival in Sweden was strongly associated with PCR positivity, aOR 11·4 (95% CI 4·6-28·1), adjusted for age group, sex, time in Sweden and reported previous antimalarial treatment (Table 3). The strong association remained when restricting the analysis to participants included at the Migrant Health Clinics, aOR 8·0 (95% CI 2·9-22·1).

**Table 3.** Risk factor analysis for PCR-positivity in migrants from Sub-Saharan Africa and in migrants attending Migrant Health Clinics.

|  | **All migrants** |  |  |  | **Migrants attending Migrant Health Clinics** |  |  |
| --- | --- | --- | --- | --- | --- | --- | --- |
|  | PCR positive, all species, n (%)<br>N=789 | OR (95% CI) | aOR (95% CI)<sup>b</sup> |  | PCR positive, all species, n (%)<br>N=480 | OR (95% CI) | aOR (95% CI)<sup>b</sup> |
| **All** | 71 (9.0) |  |  |  | 50 (10.4) |  |  |
| **Age group** |  |  |  |  |  |  |  |
| 0-5 | 14 (25.0) | 5.3 (2.4-11.3) | 4.9 (2.3-10.5) |  | 10 (21.3) | 2.7 (1.2-6.0) | 2.7 (1.2-6.1) |
| 6-12 | 14 (14.7) | 2.7 (1.1-6.5) | 2.8 (1.2-6.4) |  | 12 (14.8) | 1.8 (0.7-4.6) | 1.9 (0.7-5.1) |
| 13-18 | 11 (8.8) | 1.5 (0.7-3.3) | 1.5 (0.7-3.4) |  | 5 (5.0) | 0.5 (0.2-1.5) | 0.5 (0.2-1.4) |
| 19-39 | 22 (6.0) | 1 (ref) | 1 (ref) |  | 17 (9.0) | 1 (ref) | 1 (ref) |
| 40-59 | 9 (9.3) | 1.4 (0.6-3.1) | 1.5 (0.7-4.8) |  | 5 (11.6) | 1.3 (0.4-5.0) | 1.4 (0.3-5.3) |
| ≥60 | 1 (3.7) | 0.6 (0.1-4.9) | 0.6 (0.1-4.8) |  | 1 (7.7) | 0.8 (0.1-7.2) | 1.0 (0.1-7.0) |
| **Sex** |  |  |  |  |  |  |  |
| Male | 32 (8.4) | 1 (ref) | 1 (ref) |  | 21 (8.9) | 1 (ref) | 1 (ref) |
| Female | 39 (9.5) | 1.1 (0.7-1.9) | 1.2 (0.7-1.9) |  | 29 (11.9) | 1.4 (0.7-2.6) | 0.8 (0.4-1.4) |
| *Pregnancy, in women 16-50 years* | 1 (2.6) | 0.4 (0.1-3.2) | 0.5 (0.1-3.6) |  | 1 (20.0) | 2.3 (0.2-22.8) | 2.0 (0.2-18.1) |
| **Family member positive** | 47 (60.3) | 43.4 (19.0-98.9) | 60.2 (26.3-137.8) |  | 31 (57.4) | 28.9 (9.9-84.3) | 50.0 (20.9-119.6) |
| **Time in Sweden, days** |  |  |  |  |  |  |  |
| <90 | 38 (11.6) | 1.6 (0.7-3.9)) | 1.1 (0.4-2.6) |  | 37 (12.0) | 1.5 (0.5-4.2) | 1.2 (0.4-3.6) |
| ≥90 | 32 (7.6) | 1 (ref) | 1.1 (0.6-2.0) |  | 12 (8.6) | 1 (ref) | 1 (ref) |
| Missing data | 1 |  |  |  |  |  |  |
| **Reported previous malaria treatment, within 12 months** | 13 (19.7) | 2.8 (1.1-7.4) | 2.2 (1.1-4.4) |  | 12 (20.3) | 2.6 (0.9-7.6) | 2.4 (0.7-8.3) |
| **Uganda as country of last residence** | 53 (28.3) | 12.8 (5.8-28.3) | 11.4 (4.6-28.1) |  | 36 (27.9) | 9.3 (3.8-22.4) | 9.1 (3.2-25.8) |
| **Reported ongoing symptoms<sup>c</sup>** | 7 (8.8)<sup>c</sup> | 1.0 (0.4-2.2) | 1.1 (0.5-2.4) |  | 5 (11.1) | 1.0 (0.4-3.0) | 1.3 (0.4-3.7) |
| **Reported fever within 30 days** | 10 (9.7) | 1.3 (0.5-3.6) | 1.3 (0.6-3.0) |  | 8 (17.8) | 2.7 (0.8-8.7) | 2.6 (1.0-6.8) |
| **Anaemia<sup>a</sup>** | 20 (13.1) | 1.7 (1.0-2.9) | 1.6 (0.9-2.7) |  | 18 (18.4) | 2.5 (1.3-4.5) | 2.5 (1.3-4.9) |

Abbreviations: PCR – Polymerase Chain Reaction, OR – Odds Ratio, aOR – Adjusted Odds Ratio, CI – Confidence Interval
<sup>a</sup> Anaemia was defined according to WHO, see table 1 for definition.<sup>18</sup>
<sup>b</sup> Adjusting for adjusting for age group, sex, time in Sweden (<90 or ≥90) and reported previous antimalarial treatment within 12 months
<sup>c</sup> Headache (n=1), stomach-ache (n=0), cough (n=4), body pain (n=1) and ongoing fever (n=0).

Children (<18 years of age) had higher odds for PCR positivity compared to adults, OR 2·5 (95% CI 1·4-4·6) and even higher in 0-5 year olds OR 5·3 (95% CI 2·4-11·3) (Table 3). The PCR prevalence among children was 36/245 (14·7%), and in the migrant group relocating from Uganda, the prevalence among children was particularly high, 29/81 (35·8%) (Figure 1). The majority of the PCR positives 47/71 (66·2%), belonged to families with at least one other family member testing positive by PCR, corresponding to an OR 43·4 (95% CI 19·0-98·9) (Table 3). Interestingly, different *Plasmodium* species were often detected in families with ≥2 PCR positive members (Supplementary table 2).

**Figure 1.**
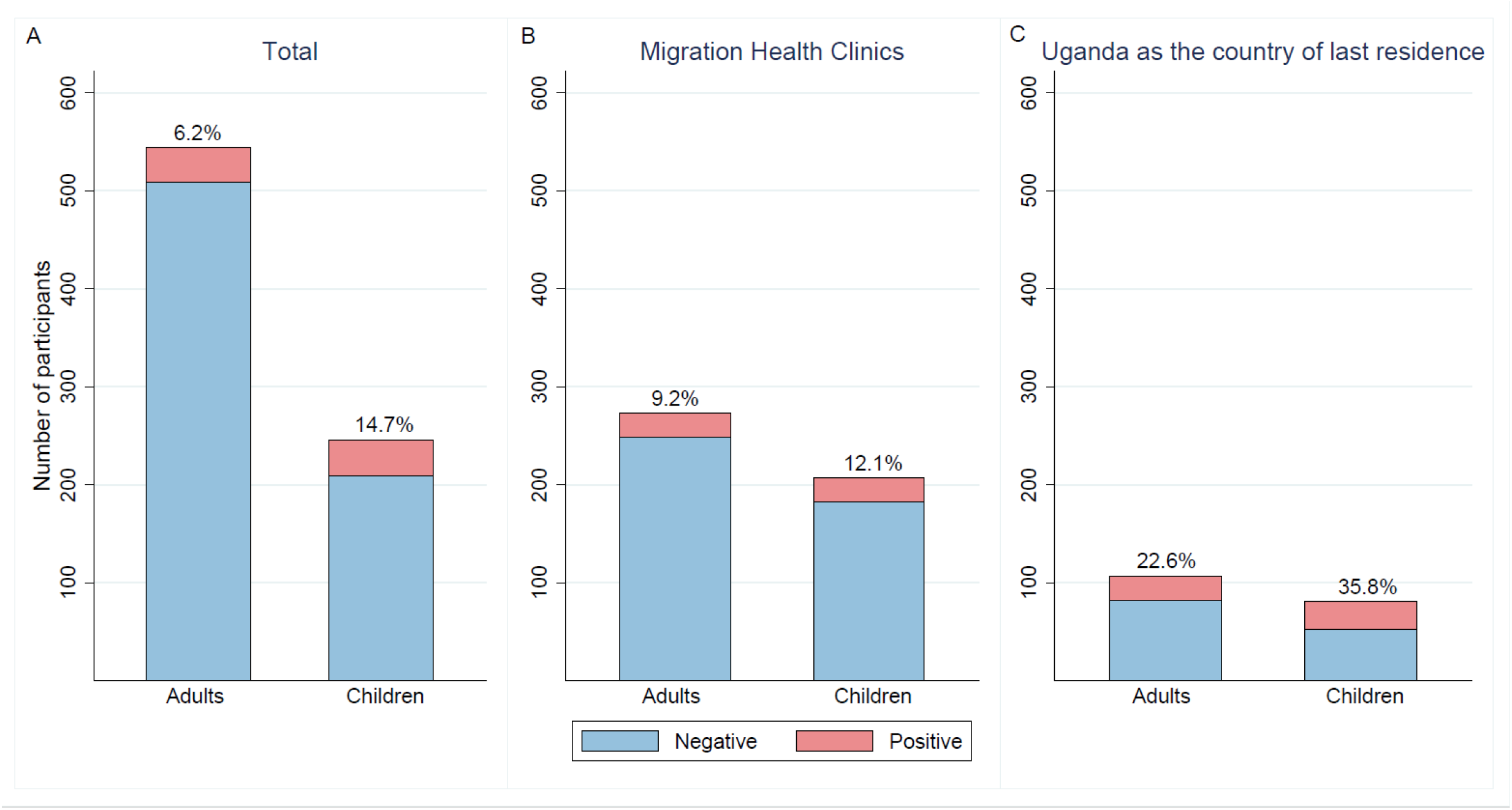
Bar graph showing malaria parasite prevalence in adult and children among participants and in the full cohort (A), participants recruited at a Migrant Health Clinic (B), and in migrants with Uganda as the country of last residence (C).

The odds for *Plasmodium* detection by PCR were similar between females and males (Table 3). Pregnancy was reported by 39/263 (14·8%) females between 16-50 years of age, and RDT and PCR positivity were detected in 1/39 (2·6%), with *P. falciparum*.

Anaemia, according to the age and sex adjusted definition by the WHO ^18^, was found in 20/71 (28.2%) of patients with PCR positivity, corresponding to an OR 1.7 (95% CI 1.0-2.9) but did not remain statistically significant in the model adjusted for age group, sex, time in Sweden and previous antimalarial treatment. No study participant had severe anaemia.

None of the participants were febrile at the time of sampling. Other ongoing symptoms were reported by 80/789 (13·0%), and 103/789 (13·1%) reported having had episodes of fever during the past month (Table 2). In the regression analysis, no statistically significant associations were found between PCR positivity and ongoing symptom or reported fever during the last month, respectively (Table 3). In participants reporting previous antimalarial treatment in the past year, 13/66 (19·7%) were PCR positive compared to 58/723 (8·0%) not reporting (p<0·01), corresponding to an OR 2·8 (95% CI 1·1-7·4) (Table 3). Previous antimalarial treatment during the past 12 months was reported by 66 participants. Of the PCR positive, time since treatment was more than 3 months in all except one PCR positive participant, reporting antimalarial treatment 15 days before participation in the study. In this case, microscopy was performed confirming asexual *P. falciparum* parasitemia.

### Duration of Plasmodium infection

Three participants tested positive after more than 365 days in Sweden, two for *P. falciparum* and one for *P. ovale*. Time lived in Sweden among PCR positive and negative individuals, respectively, is visualized in Figure 2.

**Figure 2.**
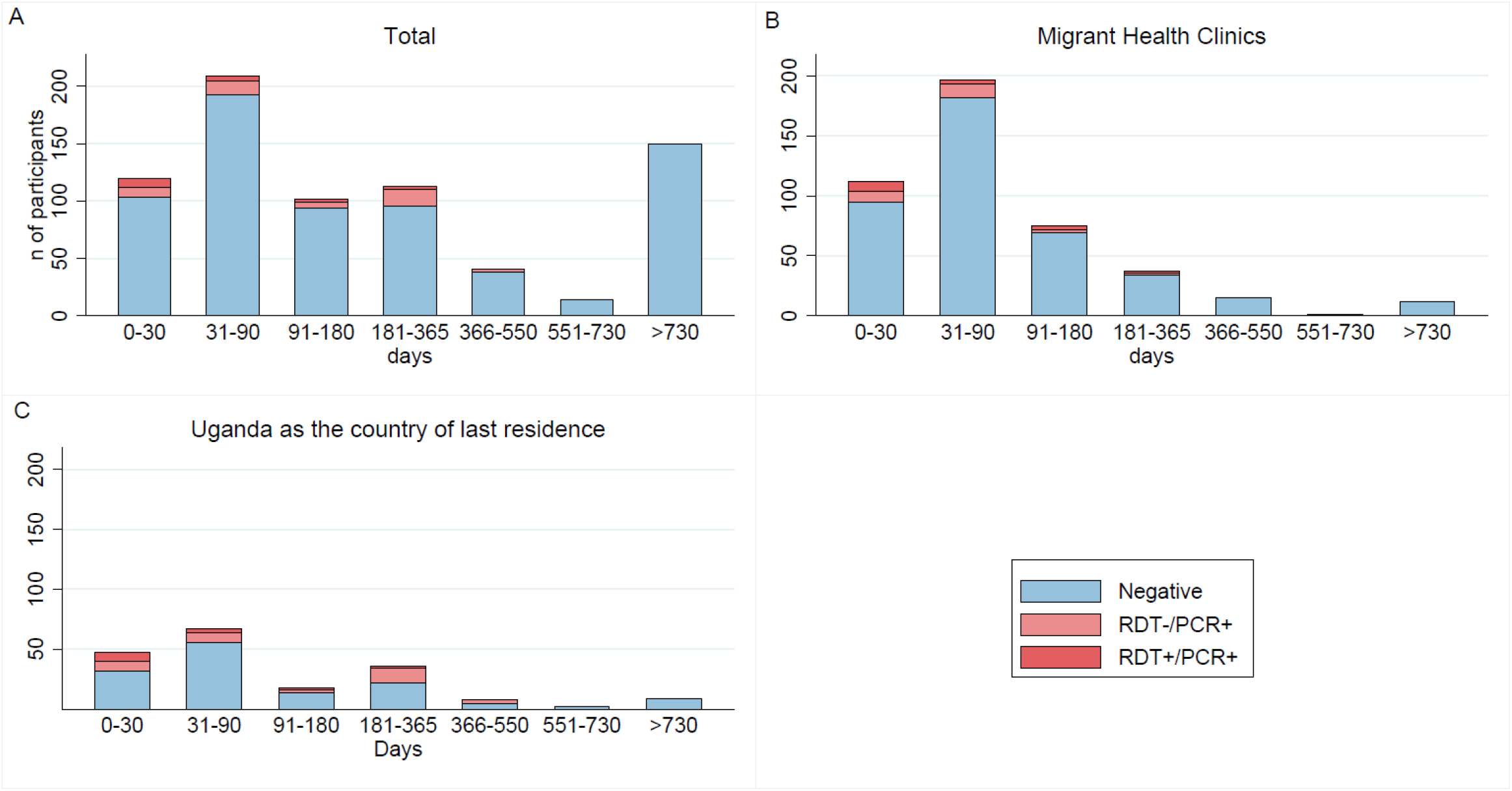
Bar graph showing malaria parasite prevalence over time of residence in Sweden, in the full cohort (A), participants recruited at a Migrant Health Clinic (B) and in migrants with Uganda as the country of last residence (C).

Comparing participants with short (<90days) and longer (≥90days) of residence in Sweden, there was no significant difference in the proportion of PCR positives, 38/329 (11·6%) vs 32/420 (7·6%), respectively (P=0·08) when comparing the full cohort; and 30/114 (26·3%) vs 23/73 (31·5%), respectively (P=0·5) in migrants with Uganda as the country of last residence.

## Discussion

In this cross-sectional study, we assessed the malaria parasite prevalence in a mixed group of migrants born in Sub-Saharan Africa and demonstrated an overall parasite prevalence of 9% in our study population of migrants with different origins and time in Sweden. In participants recruited at a Migrant Health Clinic, where screening for other infectious diseases in migrants is offered, the prevalence was 10·4%· Interestingly, most cases were found in migrants arriving from Uganda, with a prevalence of 28·3% and even higher in children, reaching 35·8%. Among the participants with a positive PCR for *Plasmodium*, the longest duration of residence in Sweden in this study population when tested was 386 days, in an individual with detected *P. falciparum*.

According to the WHO World Malaria Report 2021, DRC and Uganda are, after Nigeria, the two countries with the highest incidence of diagnosed malaria cases globally,^1^ along with a high parasite prevalence.^19,20^ In our study, the malaria parasite prevalence found in migrants with Uganda as the country of last residence was similar to the 10-35% prevalence reported from a refugee camp in Uganda.^21^ This demonstrates that migrant populations are at risk for malaria infections and may still carry the infection after arrival to a non-endemic setting. We also found PCR positive individuals among migrants arriving from Tanzania, Niger, DRC, Central African Republic, Zambia, Rwanda, Malawi, and Cameroon. However, the limited number of migrants from these countries included in our study resulted in wide confidence intervals for the estimated parasite prevalence.

Compared to microscopy and rapid diagnostic tests (RDT), molecular diagnostic methods for malaria, such as PCR have a high sensitivity, able to detect the low levels of parasitemia present in asymptomatic malaria infections.^2^ Previous studies assessing malaria prevalence among newly arrived migrants in non-endemic countries using molecular methods report prevalence ranging from 3%,^5^ to 31·8%.^4^ The malaria parasite prevalence depends largely on the origin of arriving migrants, changes over time and between migration routes, exemplified by the sharp increase of *P. vivax* malaria seen in Eritrean migrants in 2014-2015 in several European countries.^22^ During the study period, marked by the COVID-19 pandemic, DRC and Uganda were common countries of origin for migrants arriving in Sweden, and the parasite prevalence was high in this group.

The duration of *Plasmodium* infection in humans is not well described in the literature. In older data from studies of induced *P. falciparum* infections for the treatment of neurosyphilis, visible blood stage forms were reported 480 days after induction.^23^ Ashley and White^9^ have reviewed reports of prolonged blood stage infections of *P. falciparum*, however, these cases may represent the extremes of the duration of human *P. falciparum* infections. In our study, participants were included in the study irrespectively of their length of stay in a non-endemic country, which allowed for a crude estimation of the duration of infection. The longest duration of residence in Sweden was seen in an individual with *P. falciparum* with over one year since leaving an endemic area. Interestingly, the prevalence in the newly arrived (<90 days) compared to migrants with longer residence did not significantly differ, although fewer participants with longer residence were included. Also, very few participants from this high-risk group had been residing in Sweden for more than a year, limiting the analysis of the duration of infection.

*Plasmodium* infections with long incubation time, that is, blood stage infections appearing with delay after the infective mosquito bite^24^ and relapse infections occurring in *P. vivax* and *P. ovale* could potentially explain some of the PCR positive findings in participants with long duration of residence in Sweden. However, the lack of symptoms suggests a more chronic state of infection rather than a relapse prior to inclusion in the study.

As part of this study, we identified factors associated with PCR positivity. Apart from arrival from Uganda, participants aged <12 years had high odds for PCR-positivity. In addition, having a PCR positive family member was strongly associated with PCR positivity. This has also been reported in previous studies, from both endemic and non-endemic countries and increased risk in family members is likely explained by similar exposure to infective *Anopheles* mosquitoes.^25^ Therefore, testing family members for malaria should be considered around confirmed cases. Interestingly, within the identified family clusters in our study, multiple *Plasmodium* species were often found, indicating a high concomitant transmission of *P. falciparum, P. ovale* and *P. malariae* in refugee camps and other areas of residence in Uganda.

Pregnancy is a well-known risk factor for malaria infection, especially in high endemic settings, and asymptomatic malaria infections also increase the risk of adverse events during pregnancy.^10^ Due to the limited number of pregnant participants in our study, we could not evaluate the association between pregnancy and PCR positivity. Additional studies are needed to evaluate the need for *Plasmodium* screening in pregnant migrants from malaria endemic areas.

### Limitations

There are several limitations to this study. Individuals with origin in DRC and Uganda were overrepresented in the study population compared to the reported migrant population from SSA countries arriving in Sweden during the same period.^26^ The specific pathways of recruiting individuals partly contributed to selection bias, for example, our invitation letters were intentionally sent only to migrants from DRC and Uganda. In contrast, the cohort recruited in the out-patient clinic was skewed with a higher proportion of participants from Eritrea and Somalia, among whom we did not find any PCR positive. Therefore, we conducted a sensitivity analysis restricting the analysis to individuals recruited at the Migrant Health Clinics, confirming the high parasite prevalence in SSA migrants, particularly in migrants from DRC and Uganda. Apart from the letter invitations, there was despite the consecutive inclusion at the different sites no systematically collected data on the inclusion rate, and potential selection bias could therefore not be controlled for.

Since the PCR method used in this study identifies the *Plasmodium* 18S gene present in both asexual parasites and gametocytes, we could not differentiate these two forms. Gametocytes only could be present for several weeks after antimalarial treatment.^27^ In addition, PCR may be positive up to six weeks after treatment due to remaining nucleic acids.^28^ However, in the only PCR positive participant reporting antimalarial treatment within 3 months from inclusion, microscopy was performed and confirmed asexual parasitaemia. Therefore, the risk that the PCR findings in our study consists of only gametocytaemia or DNA remnants after treatment is low.

Another limitation was the cross-sectional study design, where participants were only sampled once. Low level parasitemia fluctuating close to the lower detection limit^29^ could have been missed in our study, possibly underestimating the true prevalence. However, this study was designed to resemble the setup of screening for other infectious diseases in migrants, where sampling at a single time point would be most feasible.

### Implications

Although the reintroduction of autochthonous spread of malaria in Europe is possible under optimal conditions,^30^ screening for malaria and treatment of parasite positive individuals should be considered primarily to prevent the potential negative health effects from asymptomatic malaria parasite carriage.

The short and long-term effects of low-density apparently asymptomatic *Plasmodium* infections is an understudied subject. However, asymptomatic infections may have negative effects on health, including severe complications during pregnancy, and targeting asymptomatic malaria with mass drug administration reduced all-cause mortality.^12^ This supports the need of a directed action in migrants arriving in non-endemic countries. Screening for malaria is currently not included in the national migrant health program in any European country, and further studies on long-term consequences and cost effectiveness of malaria screening should be performed.

## Conclusions

Malaria is prevalent in migrants arriving in Sweden from Sub-Saharan Africa, particularly among children. Having Uganda as the country of last residence or belonging to a family with other members testing positive for *Plasmodium*, were independently associated with PCR positivity in our study, but may differ depending on setting. Awareness of asymptomatic malaria infection in migrants from SSA among health care staff is needed. Implementation of routine screening for malaria in migrants arriving from high endemic countries, especially children, and around confirmed cases, should be considered.

## Data Availability

De-identified study data and analytic code will be made available upon request following publication and ending three years following article publication to researchers by request to the corresponding author and at the discretion of the research team.

## Acknowledgements

We express our sincere gratitude to all participants and the staff at the inclusion sites. We thank especially Ann-Mari Sjöblom, previously at Rissne Health Care Centre, for being instrumental for initiating the project, and Linnea Widman at the Biostatistics Core Facility at Karolinska Institutet for the statistical advice.

## Contributors

AW and AF conceived and designed the study. AF supervised the project. AW, RTB, IE, FL, GV, AH, SF, EH, IJ, IN, AG, SK and BT recruited participants. AW, RTB, IE, FL, GV, AH, SF, EH and IJ collected the data and performed the laboratory analyses under supervision from IB, RTB and FF. ARM, KW and OH contributed to the methodology. AW analysed the data and produced the figures. AW and AF wrote the original draft of the manuscript. AF and AW acquired funding for the study. AW, AF, ARM, IE, RTB and IB accessed and verified the data. All co-authors reviewed and edited the manuscript and were responsible for the decision to submit for publication.

---

**Supplementary table 1.**
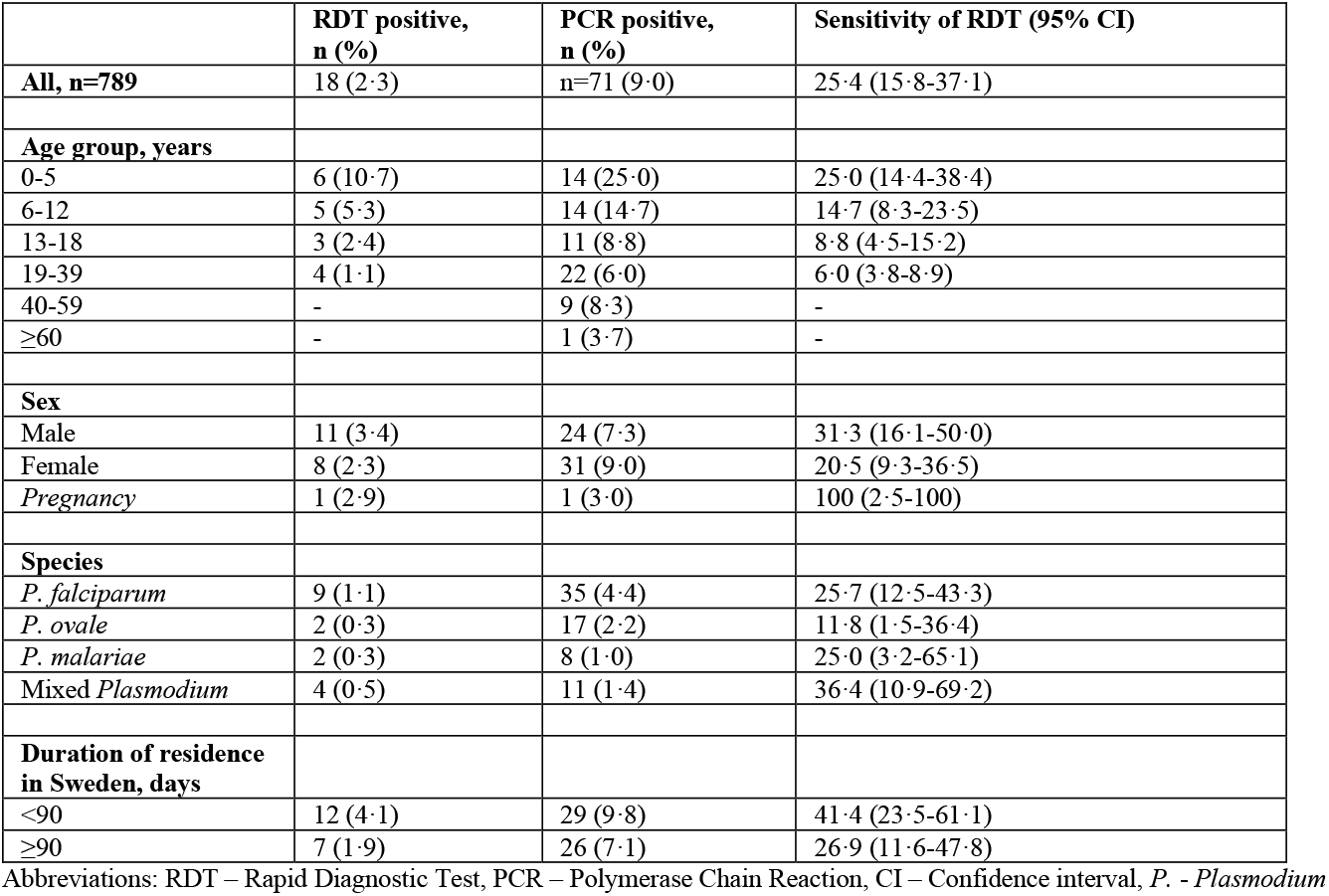
Comparison of RDT and PCR.

|  | **RDT positive,**<br>**n (%)** | **PCR positive,**<br>**n (%)** | **Sensitivity of RDT (95% CI)** |
| --- | --- | --- | --- |
| **All, n=789** | 18 (2.3) | n=71 (9.0) | 25.4 (15.8-37.1) |
| **Age group, years** |  |  |  |
| 0-5 | 6 (10.7) | 14 (25.0) | 25.0 (14.4-38.4) |
| 6-12 | 5 (5.3) | 14 (14.7) | 14.7 (8.3-23.5) |
| 13-18 | 3 (2.4) | 11 (8.8) | 8.8 (4.5-15.2) |
| 19-39 | 4 (1.1) | 22 (6.0) | 6.0 (3.8-8.9) |
| 40-59 | - | 9 (8.3) | - |
| ≥60 | - | 1 (3.7) | - |
| **Sex** |  |  |  |
| Male | 11 (3.4) | 24 (7.3) | 31.3 (16.1-50.0) |
| Female | 8 (2.3) | 31 (9.0) | 20.5 (9.3-36.5) |
| *Pregnancy* | 1 (2.9) | 1 (3.0) | 100 (2.5-100) |
| **Species** |  |  |  |
| *P. falciparum* | 9 (1.1) | 35 (4.4) | 25.7 (12.5-43.3) |
| *P. ovale* | 2 (0.3) | 17 (2.2) | 11.8 (1.5-36.4) |
| *P. malariae* | 2 (0.3) | 8 (1.0) | 25.0 (3.2-65.1) |
| *Mixed Plasmodium* | 4 (0.5) | 11 (1.4) | 36.4 (10.9-69.2) |
| **Duration of residence**<br>**in Sweden, days** |  |  |  |
| <90 | 12 (4.1) | 29 (9.8) | 41.4 (23.5-61.1) |
| ≥90 | 7 (1.9) | 26 (7.1) | 26.9 (11.6-47.8) |

Abbreviations: RDT – Rapid Diagnostic Test, PCR – Polymerase Chain Reaction, CI – Confidence interval, *P.* - *Plasmodium*

**Supplementary table 2.** Families with ≥2 PCR positive members.

| Family | Members positive (%) | Species (number of positive) |
| --- | --- | --- |
| Fam 14 | 4/5 (80) | Pf (1), Po (1), Pm (1), Pf + Pm (1) |
| Fam 15 | 5/8 (62.5) | Pf (1), Po (2), Pm (2) |
| Fam 25 | 3/5 (60.0) | Pf (3) |
| Fam 29 | 2/3 (66.7) | Po (1), Pv & Pm |
| Fam 32 | 3/3 (100) | Pf, Pf & Po & Pm (1) |
| Fam 52 | 3/3 (100) | Pf (2), Po (1) |
| Fam 54 | 3/5 (60.0) | Pf (1), Po (1), Pf & Po (1) |
| Fam 69 | 3/4 (75.0) | Pf (1), Po (1), Pm (1) |
| Fam 73 | 7/14 (50.0) | Pf (1), Po (2), Pm (2), Pf & Po (1), Pf & Po & Pm (1) |
| Fam 74 | 2/5 (40.0) | Pf (2) |
| Fam 87 | 5/5 (100) | Po (5) |
| Fam 99 | 3/15 (20.0) | Pf (2), Pf & Po (1) |

& indicates mixed *Plasmodium* infections.
Abbreviations: Pf – *Plasmodium falciparum*, Po – *Plasmodium ovale*,
Pv – *Plasmodium vivax*, Pm – *Plasmodium malariae*

**Supplementary table 3.** Risk factor analysis for PCR-positivity in migrants with Uganda as country of last residence.

|  | **Migrants with Uganda as country of last residence** |  |  |
| --- | --- | --- | --- |
|  | PCR positive, all species, n (%)<br>N=187 | OR (95% CI) | aOR (95% CI)<sup>b</sup> |
| **All** | 53 (28.3) |  |  |
| **Age group** |  |  |  |
| 0-5 | 10 (34.5) | 2.7 (0.8-8.6) | 2.8 (0.9-8.7) |
| 6-12 | 13 (41.9) | 3.7 (1.2-11.5) | 3.8 (1.2-11.7) |
| 13-18 | 8 (28.6) | 2.0 (0.7-6.0) | 2.1 (0.7-5.9) |
| 19-39 | 12 (16.4) | 1 (ref) | 1 (ref) |
| 40-59 | 9 (45.0) | 4.2 (1.6-10.7) | 3.9 (1.5-14.8) |
| ≥60 | 1 (20.0) | 1.3 (0.1-14.4) | 1.3 (0.1-14.8) |
| **Sex** |  |  |  |
| Male | 25 (30.1) | 1 (ref) | 1 (ref) |
| Female | 28 (26.9) | 0.9 (0.5-1.6) | 1.1 (0.6-2.1) |
| *Pregnancy, in women 16-50 years* | 1 (33.3) | 3.0 (0.2-47.5) | 2.7 (0.2-48.6) |
| **Family member positive** | 41 (69.5) | 22.0 (11.2-43.2) | 23.9 (9.4-60.9) |
| **Time in Sweden, days** |  |  |  |
| <90 | 30 (26.3) | 0.8 (0.3-2.3) | 0.7 (0.3-2.2) |
| ≥90 | 23 (31.5) | 1 (ref) | 1 (ref) |
| **Reported previous malaria treatment, within 12 months** | 10 (27.0) | 0.9 (0.3-3.3) | 1.1 (0.3-4.0) |
| **Uganda as country of last residence** |  |  |  |
| **Reported ongoing symptoms** | 5 (21.7)<sup>a</sup> | 0.7 (0.2-1.8) | 0.6 (0.2-2.2) |
| **Reported fever within 30 days** | 7 (29.2) | 1.3 (0.3-5.2) | 0.9 (0.3-2.6) |
| **Anaemia<sup>b</sup>** | 15 (35.7) | 1.6 (0.8-3.0) | 1.8 (0.9-3.5) |

<sup>a</sup> Headache (n=1), stomach-ache (n=1), cough (n=2), body pain (n=1) and ongoing fever (n=0)
<sup>b</sup> Anaemia was defined according to WHO, see table 1 for definition.<sup>18</sup>

